# Harnessing social media, influencers, and community-engagement to confront HIV-related stigma: implementation and evaluation of the DiME campaign

**DOI:** 10.64898/2025.11.25.25340694

**Authors:** Alyson Nunez, Milagros Wong, Marguerite Curtis, Jhoselyn Acosta, Josué Valera, David Sánchez, Yihan Shi, Ella Chun, Kristin A. Kosyluk, Jerome T. Galea, Molly F. Franke, Renato A. Errea

## Abstract

Mitigating HIV-related stigma is crucial to improving outcomes for people living with HIV (PLWH) and reaching global HIV targets. We designed and implemented a Spanish-language community-engaged social marketing campaign to address HIV-related stigma among young people in Peru, and examined the feasibility of working with influencers to disseminate campaign content. Based on formative community-engaged research with young PLWH, we developed social media content addressing common myths and stereotypes regarding HIV, encouraging empathy for PLWH, and normalizing conversations around testing, prevention and treatment. Additionally, we hired seven top Peruvian influencers (four lifestyle influencers, one physician, one reality television star, and one newscaster) to create and post content aligning with these themes from their accounts. 78 Peruvian (unpaid) micro-influencers were invited to disseminate content. All content was reviewed and approved by a community advisory board prior to posting. The campaign occurred over five months in 2024. We created and posted 49 videos to the campaign Instagram and TikTok accounts, yielding 1,475,918 views, 1,462 comments, and 63,488 likes. The campaign generated 2,206 Instagram followers and 11,800 TikTok followers, 46% of whom were in Peru. The five top posts created by the content creation agency and study team received 742,314 views, 1,108 comments, and 50,071 likes, while the five top influencer posts had a total of 2,484,934 views, 1,079 comments, and 129,258 likes. 30 unpaid micro-influencers reposted content. We received 512 direct messages inquiring about HIV services, requesting HIV-related information, or sharing personal experiences. Social media is a powerful tool for disseminating anti-stigma messaging. Collaboration with influencers to disseminate content was feasible and effective in expanding reach.

## INTRODUCTION

HIV-related stigma encompasses negative beliefs, attitudes, and subsequent behavior toward people living with HIV (PLWH). Globally, HIV-related stigma impedes prevention, testing, and treatment efforts, with negative health consequences, including preventable suffering and death^1–3^. Mitigating this stigma is, therefore, essential to improving the health of PLWH and reducing transmission.

Social media platforms, such as Instagram and TikTok, are promising tools for health promotion and education activities, particularly those targeting young people, a group highly affected by HIV-related stigma^4^. In addition to their broad accessibility and popularity, social media platforms offer a mechanism for dissemination to large audiences and opportunities for audience engagement with health messaging^5–7^. The power of social media is further potentiated by social media influencers – individuals who maintain a large and engaged audience on social media and are often trusted messengers who may affect the decisions, opinions, or behaviors of their audience.

Public health programs and research studies have used social media to disseminate educational health-related content and address misinformation surrounding infectious diseases such as COVID-19, HIV, and other sexually transmitted infections^8,9^. In recent years, some of these initiatives have also engaged social media influencers to increase audience reach. For example, one intervention included paid influencers to promote harm reduction measures for stigmatized bloodborne diseases such as HIV and Hepatitis C, finding that their participation in the campaign was a feasible strategy to promote audience reach^10^. Another study evaluated the use of influencers in a social media campaign to promote COVID-19 vaccination among the US Spanish-speaking community and found that content featuring influencers resulted in greater audience engagement compared to content that did not feature influencers^11^. While social media is increasingly gaining traction as a platform for health communication campaigns, there is limited evidence on the feasibility, implementation and evaluation of such interventions specifically targeting stigma. Here, we describe the implementation of “DiME” (Spanish for “Tell Me”), a Spanish-language,community-engaged social media campaign aimed at addressing HIV-related stigma among adolescents and young people in Lima, Peru. We evaluate the feasibility of collaborating with local social media influencers and summarize lessons learned.

## METHODS

### Study setting

Peru, the target region for the social media campaign, has an estimated HIV prevalence of 0.3-0.4%. The prevalence is notably higher in key populations such as transgender women (32%), gay, bisexual, or other men who have sex with men (10%), and female sex workers (2.4%)^12,13^. An estimated 110,000 PLWH reside in Peru, with a recent increase in the number of new HIV diagnoses in youth 18-29^14^. Social media is widely used: the country of 34 million individuals includes 10.8 million Instagram users, 3.5 million of whom are aged 18-24^6^, and over 20 million TikTok users^7^.

### Formative research to inform the campaign

We conducted formative qualitative work with a diverse group of young people, HIV advocates, and clinic providers to inform campaign content, specifically change goals, key messages, and content delivery strategies.^15^

### Social media content creation

The DiME study team consisted of individuals from Peru and the United States: an infectious disease epidemiologist, a medical doctor and HIV program manager, a nurse, a behavioral scientist and social worker, a stigma expert, a community manager and activist living with HIV, a social media content creator, and research staff. Most team members were bilingual in Spanish and English. We contracted a Peruvian content creation agency to create social media content that addressed HIV-related stigma, specifically targeting the Peruvian audience. While the agency had experience developing content related to social issues, including stigmatized infectious diseases (e.g., tuberculosis), it did not have experience creating content related to HIV or stigma. To sensitize agency team members to these topics, we provided a virtual education session on clinical information and created an explanatory report including an overview of stigma, how HIV-related stigma manifests in Lima, and a summary of formative research findings^15,16^. Guided by this information, the agency created a campaign proposal using the slogan *“Seamos una voz” (“Be the voice”)*, which focused on dispelling stereotypes and normalizing conversations around HIV while promoting empathy, not pity. Videos debunked myths (e.g., HIV and AIDS are synonymous, HIV only affects the LGBTQIA+ community, HIV is transmitted through touch, people living with HIV cannot lead “normal” lives); and educated about important concepts (e.g., undetectable = untransmissible).

The content creation agency produced a draft script of each post for review by the study team and a community advisory board made up of 15 PLWH, 4 of whom were HIV activists. Community advisory board members were diverse in terms of age (range: approximately 19 to 37 years) and sexual orientation. The study team and advisory board reviewed scripts, provided feedback, or, occasionally, recommended against producing a particular piece of content. Once produced, each piece of content underwent a second review by the study team and advisory board for final approval. The agency filmed content around Lima featuring diverse Peruvian actors and incorporated trending music, memes, sound clips, and stickers. Of the 35 total pieces of video content produced, seven did not pass the approval process and were archived.

To achieve our target posting frequency of two pieces of content per week, the study team produced 21 additional videos to supplement material created by the agency. These videos provided information about sexually transmitted infections testing and treatment in Lima, addressed stigmatizing comments lefton the DiME account, and shared testimonials from young people living with HIV (YPLWH) about their experiences with stigma.

### Collaboration with influencers

We contracted a Peruvian influencer management agency to identify and recruit influencers to participate in the campaign. We presented the agency with the stigma report, the campaign’s objectives, and the scope of work for influencer participation. The agency proposed a list of top Peruvian influencers, with diverse profiles in terms of gender, sexual orientation, and the types of content they produce. The final list of paid participating influencers included five from this list and two suggested by the study team: four LGBTQIA+ lifestyle influencers, one physician, one reality television star, and one newscaster, with follower counts ranging from 117K to 23.8M. Paid influencers were provided with information on HIV-related stigma and guidance on appropriate language to use when discussing HIV in their videos. Influencers created video scripts, which were subsequently reviewed and edited by the study team and community advisory board, before filming and posting content to their platforms. 78 micro-influencers, defined as influencers with modest followings, were identified by the management company based on perceived willingness to participate in a social campaign or past participation in campaigns for a social cause. Invited influencers had an average following count of 101,696 and a median following count of 48,000. These influencers were invited to join the campaign voluntarily by reposting content, from the DiME account or a paid influencer, to their social media pages.

### Data collection and analysis

Social media indicators (i.e., followers, views, likes, comments, saves, and shares) were monitored and recorded weekly by the study team during the campaign. Instagram professional dashboard and TikTok business suite were used to extract demographic information and performance insights (i.e., views and video watch time). Key performance indicators included: a) reach (total number of views), b) interaction (total number of likes, comments, shares, and saves), and c) engagement rate (total number of likes, shares, saves, comments, divided by the total number of views).

Top performing videos were considered to be those with the greatest total interaction at campaign end. All direct message exchanges to the DiME account on both platforms were categorically analyzed. A sentiment analysis was performed on all user comments left on campaign content. Three study team members manually categorized comments as positive, negative, or neutral, generally and with respect to the content (Supplemental Material 1). Positive comments praised the DiME initiative and expressed resonating with content, while negative comments expressed stigmatizing opinions or questioned the normalization of HIV as a chronic, treatable condition. Neutral comments included questions and answers regarding HIV, superfluous information, greetings, and ambiguous emojis. The polarity ratio was calculated as the number of positive comments divided by the number of negative comments.

After completion of the campaign, all participating paid and micro-influencers were invited to participate in a zoom interview and/or complete a questionnaire asking them about their experience participating in the campaign. Influencers provided informed oral consent prior to being interviewed over zoom. Influencers who completed the questionnaire were not asked to provide written consent but were informed participation was voluntary and anonymous.

### Campaign implementation

The DiME campaign ran from 8 July 2024 to 12 December 2024 on the Instagram and TikTok social media platforms under the account name @*dime*.*peru*. 48 videos were posted on Instagram and 49 on TikTok, with an approximate posting frequency of twice per week. For 36% of Instagram posts and 30% of TikTok posts, paid boosts were used to increase reach, with a total of approximately $564 spent across both platforms. Separately, the seven collaborating paid influencers contributed a total of 13 videos to the campaign. Influencers posted content to their own social media accounts and directed their followers to the DiME account. Six of these videos were posted in collaboration with the DiME account, appearing simultaneously on the influencer’s page and the DiME Instagram account (collaborations were not possible on TikTok during the study period). The DiME community manager monitored the comment sections on all videos and direct messages to both accounts and provided timely responses to questions, misinformation, and requests for support.

## RESULTS

### Key performance indicators

During the five-month campaign, the DiME account gained 2,206 followers on Instagram and 11,800 followers on TikTok. According to platform analytics obtained in December 2024 for the last month of the campaign, approximately 46% of followers had Peru-based accounts, 64% were male, and 56% were aged 18-34. Median reach per post after the first four weeks of the campaign was 470 on Instagram and 994 on TikTok and increased to 4,011 and 3,203, respectively, at campaign end. Median interactions per post increased from 30 on Instagram and 54 on TikTok within the first four weeks, to 86 and 110, respectively, at campaign end. Our overall engagement rate at campaign end was 1.65% on Instagram and 6.89% on TikTok. Influencer content generated over 2.8M views and 160K interactions. Metrics reported in Table 1 reflect numbers though the campaign end in December 2024.

**Table 1.**
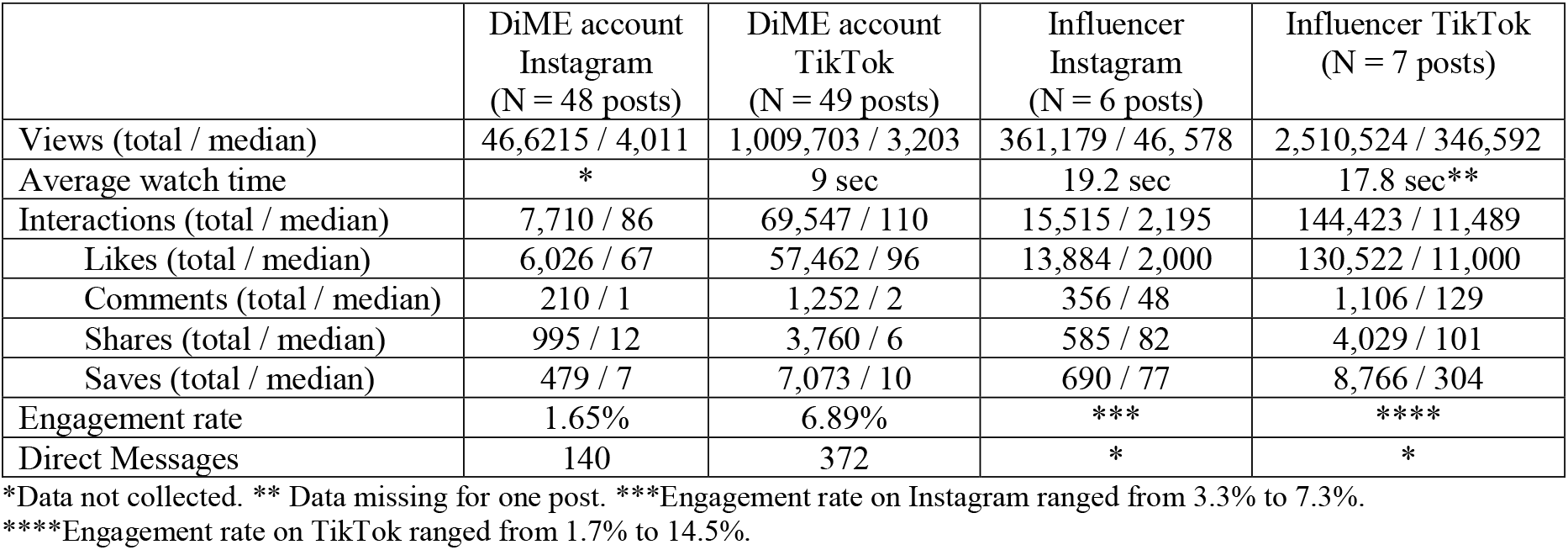
Overall metrics for DiME and influencer content (8 July 2024 – 12 Dec 2024)

|  | DiME account<br>Instagram<br>(N = 48 posts) | DiME account<br>TikTok<br>(N = 49 posts) | Influencer<br>Instagram<br>(N = 6 posts) | Influencer TikTok<br>(N = 7 posts) |
| --- | --- | --- | --- | --- |
| Views (total / median) | 46,6215 / 4,011 | 1,009,703 / 3,203 | 361,179 / 46,578 | 2,510,524 / 346,592 |
| Average watch time | * | 9 sec | 19.2 sec | 17.8 sec** |
| Interactions (total / median) | 7,710 / 86 | 69,547 / 110 | 15,515 / 2,195 | 144,423 / 11,489 |
| Likes (total / median) | 6,026 / 67 | 57,462 / 96 | 13,884 / 2,000 | 130,522 / 11,000 |
| Comments (total / median) | 210 / 1 | 1,252 / 2 | 356 / 48 | 1,106 / 129 |
| Shares (total / median) | 995 / 12 | 3,760 / 6 | 585 / 82 | 4,029 / 101 |
| Saves (total / median) | 479 / 7 | 7,073 / 10 | 690 / 77 | 8,766 / 304 |
| Engagement rate | 1.65% | 6.89% | *** | **** |
| Direct Messages | 140 | 372 | * | * |

\*Data not collected. \*\* Data missing for one post. \*\*\*Engagement rate on Instagram ranged from 3.3% to 7.3%.
\*\*\*\*Engagement rate on TikTok ranged from 1.7% to 14.5%.

### Sentiment analysis

Of the 1,462 total comments received on all DiME posts, 684 (47%) were positive, 546 (37%) were neutral, and 232 (16%) were negative. A small percentage of comments 6%) conveyed anti-stigma messaging using hostility or negativity towards stigmatizing opinions. We classified these as positive if they endorsed the content messaging. The resulting polarity ratio (positive: negative) was 2.95: 1, indicating nearly three times as many positive to negative comments. DiME account followers frequently responded to negative comments with their own messages of education:

### (Regarding discrimination in the workplace)

*User 1: At a private company you can hire based on internal policy. So start a company and hire people with your own policies. Period*.

*User 2: No internal policy is above the constitution, and discrimination on the basis of disease is a crime*.

### (Regarding serodiscordant couples)

*User 1: All the same, use protection. Don’t make [HIV] normal just because it’s your opinion*.

*User 2: You’re uninformed. If you research and ask specialists, everything they’re saying is 100% correct. Undetectable equals untransmissible even if you have sex with someone who doesn’t have it*.

Influencers received a total of 1,462 comments on content produced for the DiME campaign, 64.8% of which were positive. Many of their followers congratulated them for informing their audiences about HIV: *“More influencers like you who don’t just entertain but also inform*.*” or “I learned more in two minutes with Josi than in five years of school!”*

### Direct messages

We received 512 direct messages to our accounts corresponding to the following categories: requests for HIV and STI testing locations (59.8%), questions regarding HIV testing protocols, transmission, and treatment (12.6%), sharing of personal experiences (1.6%), and other (26%). The other classification was used for messages praising the DiME campaign, messages asking where DiME was physically located, and greetings without follow-up, among others. Messages originated from 37 localities in Lima, 16 Peruvian provinces, and seven Latin American countries. The DiME community manager directed users to the appropriate information and/or provided guidance and encouragement. Illustrative messages are included in Table 2.

**Table 2.** Illustrative direct messages by category.

|  |  |
| --- | --- |
| Requests for HIV and STI testing locations | *Hi, I saw your video about syphilis testing and I wanted to know where I could get a free test in Lima. Thank you so much.*<br>*Hi, I saw you on TikTok. How can I get an HIV test? I'm from San Juan de Lurigancho.* |
| Questions regarding HIV testing protocols | *I'm not 18 yet, can I go alone as a minor? It's just that I don't want my parents to find out, I'd feel horrible. Seriously thank you, I don't know who's behind the screen but you're giving me peace.*<br>*Hi, could you help me with how long I should wait to take an HIV test? I asked and they offered me a 4<sup>th</sup> generation immunochromatographic test, is that one good? I've been waiting three weeks and I'm starting to have symptoms and I don't know if it's because of that or the stress of not knowing.* |
| Questions regarding HIV transmission | *I want to know if a person on treatment and undetectable can get someone pregnant.*<br>*Hi, I just wanted to know if someone who's seropositive can transmit HIV. I had a seropositive partner and at first when they told me I was scared, and I had this fear of getting it. Now we're not together and I'm scared they gave it to me. During the relationship I got tested every three months.*<br>*My question is if you can also get it from a tattoo (I got a tattoo and I didn't pay attention to the needle) and I'm scared.* |
| Questions regarding HIV treatment or PrEP | *So if I have HIV and the other person too, can we both take PREP?*<br>*I got the diagnosis and I'm wondering where I can start treatment as soon as possible.* |
| Sharing of personal experiences | *Hi, I'm new with HIV and I'm not taking it as well as everyone else :( I don't know if I'll have a partner or friends or if I'll have a good life.*<br>*I want to file a legal complaint. How can I do that, can you advise me? I'm HIV+ and my partner wants to tell everyone.*<br>*I don't know who I'm lucky enough to be talking to but it's really helpful to talk with someone since I'm dealing with this alone.*<br>*Hi, this is an awesome initiative. I wish I had this information a long time ago. Someone close to me died almost three years ago and I couldn't help him. God-willing someday I can help someone else in the name of my brother.* |
| Other | *I love this, really it's so nice to make this visible so people get a little more informed and to help them see that with treatment people can live normal lives. If you're ever thinking about a reel or podcast I'd love to participate.*<br>*Where are you located? Because I'm HIV+ and I had to flee Venezuela because I couldn't access treatment there. I'm backpacking until I can get settled and get my treatment. I wanted to know if you help in these cases.* |

### Top performing content

Metrics for top-performing content are shown in Table 3. Of the content created by the agency and study team, the top post depicted actors representing a young male same-sex couple in a sero-discordant relationship. They were filmed in Lima dispelling myths about sexual relationships while living with HIV and encouraging their peers to examine preconceived notions. This video was posted on 8 November and went viral, spreading rapidly to reach a wide audience within a short period of time. It received 388,276 views, 17 times the average number of views for DiME content, and 48,646 interactions, 80 times the average interactions. Other top performing posts, as defined by number of total interactions, dealt with rights in the workplace for YPLWH, the difference between HIV and AIDS, and the stereotype that YPLWH cannot live like their peers. These videos all received more than 65K views and 2,000 interactions.

**Table 3.** Metrics for top-performing content created by agency and study team and by influencers.

| Post content | Views | Total interactions | Likes | Comments | Saves | Shares | Engagement rate | % positive comments | % neutral comments | % negative comments |
| --- | --- | --- | --- | --- | --- | --- | --- | --- | --- | --- |
| **Agency and study team** |  |  |  |  |  |  |  |  |  |  |
| Same sex sero-discordant relationship | 388,276 | 48,643 | 39,553 | 500 | 5,487 | 3,103 | 12.53% | 54.2% | 38.8% | 7% |
| Account of YPLWH not hired due to HIV | 95,819 | 3,809 | 3,034 | 437 | 226 | 112 | 4% | 34.1% | 37.5 | 28.4% |
| Rights of YPLWH in the workplace | 69,259 | 3,540 | 3,055 | 75 | 259 | 151 | 5.1% | 61.3% | 18.7% | 20% |
| Doctor explaining HIV vs. AIDS | 116,474 | 2,912 | 2,626 | 34 | 158 | 94 | 2.5% | 38.2% | 50% | 11.8% |
| A person finds out their neighbor is living with HIV | 72,486 | 2,131 | 1,803 | 62 | 162 | 104 | 2.9% | 45.2% | 45.1% | 9.7% |
| **Influencer** |  |  |  |  |  |  |  |  |  |  |
| LGBTQ+ male, lifestyle influencer, teaches a “class” dispelling HIV-related myths | 593,000 | 57,149 | 52,000 | 413 | 3,639 | 1,097 | 9.6% | 68.3% | 28.1% | 3.6% |
| Male newscaster explains HIV can affect anyone | 676,000 | 39,598 | 34,000 | 171 | 3,062 | 2,365 | 5.9% | 48% | 47.3% | 4.7% |
| Transwoman, lifestyle influencer, debunks HIV-related stereotypes | 180,000 | 26,083 | 24,000 | 282 | 1,438 | 363 | 14.5% | 69.5% | 24.5% | 6% |
| Female reality TV star discusses empathy | 689,342 | 11,489 | 11,000 | 84 | 304 | 101 | 1.7% | 52.4% | 44% | 3.6% |
| Female reality TV star dispels myths about HIV | 346,592 | 8,743 | 8,258 | 129 | 289 | 67 | 2.5% | 45.7% | 51.2% | 3.1% |

The five top-performing influencer pieces were all posted on the TikTok platform and included videos created by lifestyle influencer members of the LGBTQIA+ community, a male social media newscaster, and a female reality television celebrity. These videos all received at least 180K views and 8,000 interactions.

Figure 1 provides a comparison between reach and interaction metrics for the top-performing videos created by the agency and study team and by influencers. Influencer numbers were greater for all categories with the exception of comments received.

**Figure 1.**
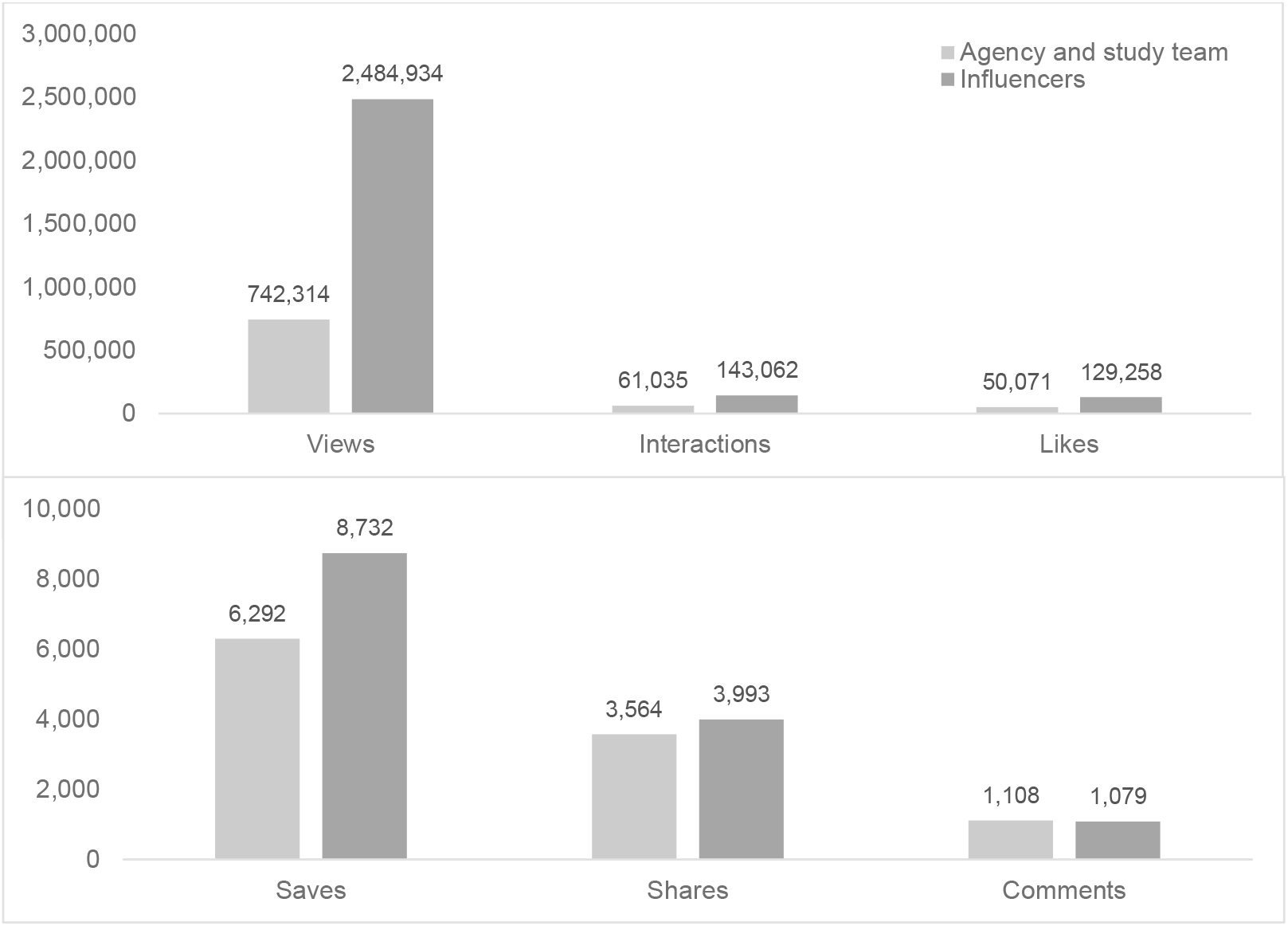
Comparison of reach and interaction indicators for top-performing content

### Micro-influencers

30 unpaid micro-influencers (38% of total invited) shared a total of 27 and 9 posts on Instagram, created by the agency and influencers, respectively. Three influencers created their own content about HIV-related stigma and directed their followers to the DiME account. Seven micro-influencers that were not part of the project also reposted campaign content.

### Influencer experience participating in the campaign

Three paid influencers and two micro-influencers responded to our post campaign survey. Three reported prior experience participating in campaigns to raise awareness about social issues but none had experience with HIV-related campaigns. All reported accepting the invitation to participate in the campaign because it aligned with their personal values. These influencers reported mostly positive audience reactions to campaign content. Two indicated that participating in the campaign positively affected their image on social media, while three mentioned the campaign had neither a positive nor negative effect on their image. All reported willingness to participate in future HIV-related social media campaigns.

We carried out in-depth interviews with two micro-influencers who shared content: a comedian and theater professor with 6K followers and a fashion and skincare creator with 66K followers. Both praised thecampaign and explained that using their platforms to amplify social causes was important to them. They chose to follow the DiME account to add to their existing knowledge base about HIV, though one mentioned that receiving more information about the campaign, HIV, and stigma would have been useful. Both highlighted the importance of consistent posting frequency in maintaining account momentum.

## Discussion

Social media has become an effective and economical tool for distributing public health information^17–19^; however, campaigns explicitly seeking to address stigma are lacking. The DiME social media campaign sought to reduce HIV-related stigmatizing opinions and behaviors in the general population and self-stigma in YPLWH in Peru by addressing stereotypes and promoting empathy through educational messages, interviews, and testimonials. A diverse group of actors and spokespeople from the local community established the trust necessary to build a committed following, and content filmed in Lima enabled the audience to connect with the campaign messaging, identifying with the local context. Videos created and shared by influencers expanded campaign reach while helping to normalize the discussion of HIV. Though this report focused on campaign implementation, other analyses will examine whether campaign content and/or exposure to the campaign reduced stigmatizing opinions among youth generally and among those living with HIV.

Our experience suggests that collaborating with paid mega- and unpaid micro-influencers on HIV-related social media campaigns is both feasible and advantageous. Working with a local influencer management agency allowed us to efficiently identify and recruit strategic personalities willing to participate. Reach and engagement with influencer-produced content was substantially greater than with content developed by a content creation agency, in collaboration with the DiME team. Micro-influencers’ sharing of DiME content further increased campaign visibility. These observations align with findings from previous public health campaigns that suggest influencers can positively communicate health information and increase engagement among target audiences^11,20–22^. In Latin America, conversation around HIV continues to be taboo, and reaching large audiences who are not routinely exposed to HIV-related information is of utmost importance for stigma reduction^23^. We found that influencers were a highly effective means of reaching a wider and more diverse audience than could otherwise be achieved. Influencers were, in general, receptive to instruction for respectful presentation of HIV-related content and their agreement to address these topics, despite potential negative comments, is a step toward broader public interest in HIV education. However, it is important that content creators receive training on HIV and HIV-related stigma prior to production in order to ensure that the information they share is communicated in a way that reduces stigma rather than reinforces it, while maintaining the creator’s personal style.

Establishing trust between a campaign and its audience should be a central objective of social media initiatives^24^. Previous research has shown that digital platforms can serve as trusted spaces where YPLWH have the confidence to discuss stigma and seek support^25–27^. At the end of the five-month campaign, we had built a strong community on both platforms where followers shared personal experiences and asked for advice through direct messages or public comments. The role of the community manager was important for obtaining this trust. This position was filled by a person living with HIV with experience in peer counseling and HIV communication, bringing lived experience and promoting authentic connection with the audience. This strengthened the campaign’s credibility and enhanced the relatability of interactions with users. Preserving a balance between quantitative reach and qualitative human connection is crucial to maintaining the relational dimension that enables stigma reduction to occur. The process of trust-building took time. For the first half of our campaign period, reach and engagement were much lower than at campaign end.

As such, determining the appropriate duration for public health social media campaigns should account for the time needed to cultivate an active and engaged following. Organizations should also have a transition plan or taper phase for campaign end as once the account becomes a trusted source of information followers expect constant engagement.

Among the lessons learned was the importance of developing content weeks in advance of the target posting data, as the nature of HIV and stigma information requires adequate time for thoughtful input regarding how content is perceived. Video first drafts were more effective than written scripts at conveying appropriate tone and visual appeal. Furthermore, language used in campaign content is critical^28^. We sought to strike a balance between destigmatizing language and that which is understood by a lay population. For example, in Spanish ‘to infect’ (contagiar) is more common colloquially than the preferred term ‘to transmit’ (transmitir). Though YPLWH may perceive the term ‘infect’ to be derogatory, ‘transmit’ is less commonly used and may hinder comprehension or relatability. These nuances underscore the importance of convening advisory committees of YPLWH and young people from the target population (i.e., those not known to be living with HIV) to help strike the balance of comprehension, relatability, and respect. If possible, YPLWH should accompany video recording sessions to ensure that the inadvertent use of stigmatizing language can be corrected in real time and that recorded situations mirror realistic interactions to the greatest extent possible.

Though a strength of this work was our constant communication and collaboration with an advisory committee of YPLWH, this lengthened the content creation process which led to a relatively low posting frequency of two videos per week. Social media algorithms reward high posting frequencies ^29,30^. We chose not to create content in advance in order to identify opportunities to incorporate social media trends into DiME content. However, the length of the content validation process meant that trends sometimes became obsolete by the time content was approved. Additionally, exact reach and engagement are difficult to assess. Stigma may hinder young people from following an HIV-related account or interacting with its content, although they have viewed it. This raises the possibility that engagement is greater than suggested by the metrics provided by campaign platforms. The occasional use of paid boosts also makes it difficult to distinguish between organic versus paid reach and engagement. Finally, we recognize a lack of metrics to assess the qualitative impact of community management (e.g., changes in perceptions/opinions). This paves the way for future studies that measure the narrative transformation of social media users.

Social media has the power to educate and cultivate empathy^31^, and the potential to positively influence behavior among adolescents and young adults who are just beginning to navigate the healthcare system independently^32,33^. The DiME campaign utilized educational videos, testimonials from YPLWH, and influencer content to bring HIV education to this population in an accessible and engaging way. We found TikTok to be the more effective platform for an anti-stigma campaign aimed at young people; our TikTok engagement rate of 6.89% is higher than the average 2.3% reported for the education sector on this platform^34^. However, both DiME accounts have continued to grow since campaign end and have garnered a place in the HIV space in Peru, with tags from other organizations’ HIV-related content. Direct messages indicate there is a critical need for messaging to reduce HIV-related discrimination. Future campaigns should consider targeting older populations or specific groups, such as healthcare providers, from whom stigma and discrimination may be especially harmful^35,36^.

## Conclusion

The DiME social media campaign to reduce HIV-related stigma challenged stereotypes about HIV and promoted empathy towards PLWH through short, engaging videos tailored to young people in Lima, Peru.

Collaboration with popular local influencers to create and disseminate content was a novel approach in this context and served to increase campaign reach and engagement. Influencers received mostly positive feedback from their followers and were willing to participate in future HIV-related campaigns. The DiME campaign is one of the first to explicitly address HIV-related stigma via a community-based social media campaign, adding to the growing evidence that digital platforms can serve as powerful tools for stigma reduction across languages, cultures, and health conditions. Future work should explore how social media can be used to increase linkage to care.

## Data Availability

All data produced in the present study are available upon reasonable request to the authors.

## Authorship

Conceptualization: MF, RE, JG, KK, MW, AN

Data curation: MC, MW, YS, EC, AN

Formal analysis: MC, MW, AN

Funding acquisition: MF, RE, JG, KK

Investigation: all authors

Methodology: MF, RE, JG, KK, MW

Project administration: MW, JA, DS, JV

Supervision: MF, RE

Writing – Original draft preparation: AN, MC

Writing – Review and editing: all authors

All authors have read and approved the final manuscript.

## Acknowledgements

The authors are indebted to the Mezqla Digital Agency for the production of campaign content and to the Influencers Connect agency for contacting and hiring influencers to participate in the DiME campaign.

We would also like to express our gratitude to the Socios en Salud Youth Advisory Board and the DiME Advisory Committee for their guidance throughout our project.

