## Supplemental Material 1 for "Harnessing social media, influencers, and community-engagement to confront HIV-related stigma: implementation and evaluation of the DiME campaign"

Supplemental Material – Sentiment Analysis Codebook

| Positive |  |
| --- | --- |
| *Praise* | Supporting message  Congratulating campaign  Expressing gratitude for information |
| *Agreeing* | Amplifying facts from video  Adding factual info  Agreeing with positive comments from DiME or others |
| *Refuting* | Refuting stigmatizing ideas with facts |
| *Resonating* | Giving credence to the message |
| *Lack of information* | Lamenting lack of understanding or information  Telling people to get informed |
| *Emojis* | Using supportive emojis (e.g. hearts, fire, clapping) |
| Negative |  |
| *Stigmatizing* | Voicing stigmatizing opinion  Amplifying stigmatizing opinion  Suggesting influencer has HIV |
| *Skepticism* | Expressing skepticism that information is true |
| *Misinformation* | Providing misinformation  Responding to a user comment with misinformation |
| *Refuting* | Refuting correct information in comments |
| *Reinforcing* | Agreeing with someone’s negative comments |
| *Criticizing* | Criticizing how info is presented |
| *Bullying* | Taunting others |
| *Emojis* | Using stigmatizing emojis |
| Neutral |  |
| *Superfluous* | Unrelated or unverified information  Commenting on aspects of video unrelated to HIV Tagging other users  Explaining personal issues |
| *Asking* | Asking genuine questions |
| *Uninterpretable* | Using ambiguous emojis  Generally uninterpretable information |
| *Greetings* | Saying hello to DiME account |
| *Suggestions* | Suggesting ideas for future content or ways of presenting information |
| *Engaging* | Interacting with HIV topics unrelated to video  Explaining how they would feel |
| Supportive though negative |  |
| *Insulting* | Supporting anti-stigma message through hostility or negativity |
